# Immunogenicity of COVID-19 Tozinameran Vaccination in Patients on Chronic Dialysis

**DOI:** 10.1101/2021.03.31.21254683

**Authors:** Eva Schrezenmeier, Leon Bergfeld, David Hillus, Joerg-Detlev Lippert, Ulrike Weber, Pinkus Tober-Lau, Irmgard Landgraf, Tatjana Schwarz, Kai Kappert, Ana-Luisa Stefanski, Arne Sattler, Katja Kotsch, Thomas Doerner, Leif Erik Sander, Klemens Budde, Fabian Halleck, Florian Kurth, Victor Max Corman, Mira Choi

## Abstract

**Objective:** Patients with kidney failure have notoriously weak responses to common vaccines. Thus, immunogenicity of novel SARS-CoV-2 vaccines might be impaired in this group. To determine immunogenicity of SARS-CoV-2 vaccination in patients with chronic dialysis, we analyzed the humoral and T-cell response after two doses of mRNA vaccine Tozinameran (BNT162b2 BioNTech/Pfizer).

**Design, Settings, and Participants:** This observational study included 43 patients on dialysis before vaccination with two doses of Tozinameran 21 days apart. Overall, 36 patients completed the observation period. Serum samples were analyzed by SARS-CoV-2 specific antibodies ∼1 and ∼3-4 weeks after the second vaccination. In addition, SARS-CoV-2-specific T-cell responses were assessed at the later time point by an interferon-gamma release assay (IGRA). Outcomes at later timepoints were compared to a group of 44 elderly patients with no dialysis after immunization with Tozinameran.

**Exposures:** Blood drawings during regular laboratory routine assessment right before start of dialysis therapy or at the time of vaccination and at follow-up study visits.

**Main Outcomes and Measures:** Assessment of immunogenicity after vaccination against SARS-CoV-2 in patients on and without dialysis.

**Results:** Median age of patients on chronic dialysis was 74.0 years (IQR 66.0, 82.0). The proportion of males was higher (69.4%) than females. Only 20/36 patients (55.6%, 95%CI: 38.29-71.67) developed SARS-CoV-2-IgG antibodies at first sampling, whereas 32/36 patients (88.9%, 95%CI:73.00-96.38) demonstrated seropositivity at the second sampling. Seroconversion rates and antibody titers were significantly lower compared to a cohort of vaccinees with similar age but no chronic dialysis (>90% seropositivity). SARS-CoV-2-specific T-cell responses 3 weeks after second vaccination were detected in 21/31 vaccinated dialysis patients (67.7%, 95%CI: 48.53-82.68) compared to 42/44 (93.3%, 95%CI: 76.49-98.84) in controls of similar age.

**Conclusion and Relevance:** Patients on dialysis demonstrate a delayed, but robust immune response three weeks after the second dose, which indicates effective vaccination of this vulnerable group. However, the lower immunogenicity of Tozinameran in these patients needs further attention to develop potential countermeasures such as an additional booster vaccination.

## Introduction

The coronavirus disease 2019 (COVID-19) pandemic has led to an urgent need for effective strategies, in particular for patients with kidney failure (KF), who have a high mortality [1]. The COVID-19 vaccine Tozinameran (BioNTech/Pfizer) showed protection from 12 days after the first dose [2] and was demonstrated to be 95% effective in preventing COVID-19 [3]. Robust SARS-CoV-2 antibody responses ≥7 days after the second dose were detected [4]. However, patients with KF have an impaired response to vaccination [5] and were excluded from previous trials. We investigated immunogenicity to COVID-19 vaccination with Tozinameran in 43 patients with chronic kidney disease (CKD) stage 5.

## Methods

Serum samples of vaccinated dialysis patients (Table 1, Supplement for cohort details) were analyzed for anti-SARS-CoV-2 antibodies ∼1 and ∼3-4 weeks after the second dose by anti-SARS-CoV-2-S1 IgG/IgA ELISA (Euroimmun, Lübeck, Germany). At the later timepoint, sero-response was compared to a group of 44 Tozinameran vaccinated persons of similar age but not on dialysis and to 18 unvaccinated control patients on dialysis. Comparison was based on a solid phase immunoassay (SeraSpot®Anti-SARS-CoV-2 IgG, Seramun Diagnostica, Heidesee, Germany) using four antigens, full spike (fS), spike S1 domain, spike receptor binding domain (RBD), as part of the vaccine, and the nucleocapsid antigen (N), to discriminate between vaccine and SARS-CoV-2 infection induced response. Anti-SARS-CoV-2-N-IgG positive patients were excluded from further analysis. Potential neutralizing antibodies were tested by a surrogate virus neutralization test (sNT, cPass, Medac, Wedel, Germany) [6]. SARS-CoV-2-specific T cell responses were determined by an interferon-gamma release assay (IGRA) (AG, Lübeck, Germany). Details of laboratory methods and statistical analyses are given in the supplement.

**Table 1.** Patient characteristics.

| **Age** | Vaccinated Dialysis Patients<br>(*n = 36 patients on dialysis*) | Vaccinated Non-Dialysis Controls<br>(*n = 44 control patients without dialysis*) | Unvaccinated Dialysis Patients<br>(*n = 18 patients on dialysis*) |
| --- | --- | --- | --- |
| Median years [IQR] | 74.0 [66.0, 82.0] | 80.0 [75.75, 82.25] | 63 [45.25, 74.5] |
| Under 50 | 1 | 4 | 6 |
| Between 51-59 | 5 | 1 | 2 |
| Between 60-69 | 10 | 3 | 4 |
| > 70 | 20 | 36 | 6 |
| Male | 25 | 14 | 12 |
| Female | 11 | 30 | 6 |
| **Renal replacement therapy** |  |  |  |
| Hemodialysis | 34 |  | 17 |
| Peritoneal dialysis | 2 |  | 1 |
| Years since start of renal replacement therapy |  |  |  |
| Median [IQR] | 5 [2, 9] |  | 2 [1,3] |
| **Co-Morbidities** |  |  |  |
| Hypertension | 29 | 22 |  |
| Diabetes mellitus | 16 | 9 | 2 |
| Obesity (BMI>30) | 13 | 5 |  |
| Malignancy (recent or history of) | 4 | 8 | 2 |

## Results

Three out of 43 dialysis patients developed symptoms of COVID-19 and were SARS-CoV-2 positive, as determined by PCR within 11, 14, and 16 days after one dose of Tozinameran, of whom one patient died from COVID-19. A further three patients were lost to follow up and one patient died of non-COVID-19 reasons (**eFigure 1 in Supplements**). Blood samples taken ∼1 week and ∼3-4 weeks after the second vaccination were available for 36 patients in the dialysis cohort. Patients had a median age of 74.0 years (IQR 66.0, 82.0) with a female to male ratio of 11:25. The vaccinated non-dialysis cohort had a median age of 80.0 years (IQR 75.75, 82.25), with a female to male ratio of 30:14. Patient characteristics are summarized in **Table 1**. One week after the second dose of Tozinameran, 20/36 sera of dialysis patients (55.56%; 95%CI: 38.29-71.67) were reactive for anti-SARS-CoV-2-IgG (**Figure 1A, B)**. Antibody response rate increased to 32/36 patients (88.9%; 95%CI: 73.0-96.4) within three weeks after the second dose and 33/36 sera (91.67%; 95%CI: 76.41-97.82) were also reactive for anti-SARS-CoV-2-IgA (**eFigure 2 in Supplements**). Neutralizing capacity of antibodies was detected in 28/36 (77.78% 95%CI: 60.42-89.28) sera using sNT (**Figure 1C**).

**Figure 1:**
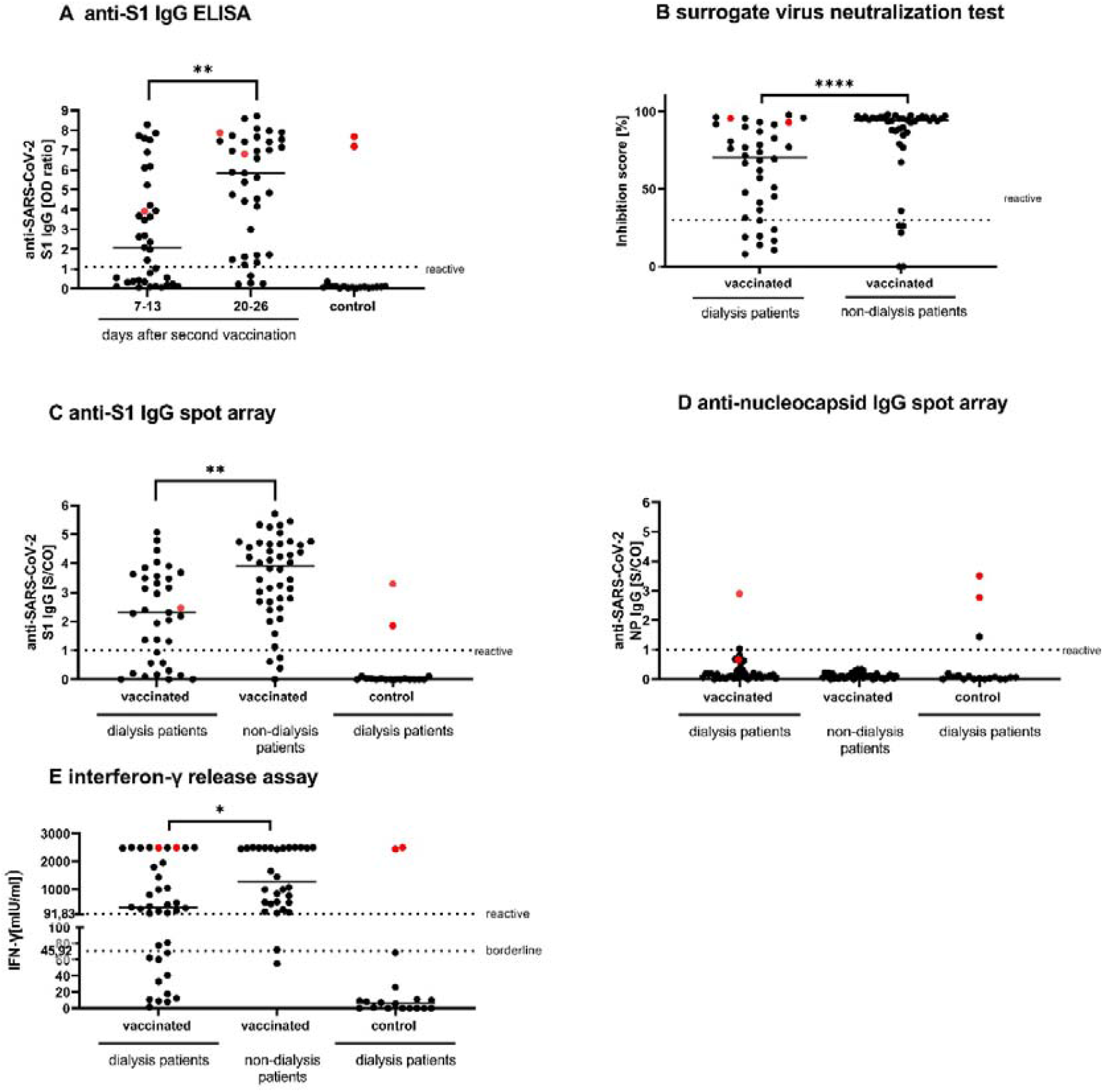
SARS-CoV-2 specific IgG and IFN-γ responses in vaccinated dialysis and non-dialysis patients. (**A**) Anti-SARS-CoV-2 S1 IgG were measured in serum of dialysis patients on day 7-13 (n=37) and day 20-26 (n=38) after a second dose of Tozinameran (BNT1621b2) and unvaccinated dialysis patients (n=18) using EUROIMMUN ELISA. Anti-S1 IgG (**B**), a surrogate virus neutralization test (**C**) as well as anti-nucleocapsid IgG (**D**) were measured 20-26 days after the second vaccination in dialysis patients, vaccinated individuals of similar age (n=44), and unvaccinated dialysis patients (n=18) utilizing the SeraSpot Anti-SARS-CoV-2 IgG assay and Medac cPass. One vaccinated dialysis patient was excluded as the internal assay control failed. **(E)** Whole blood from vaccinated and unvaccinated dialysis patients and non-dialysis individuals was stimulated *ex vivo* with a SARS-CoV-2 S1 peptide pool and IFN-γ concentration in the supernatant was measured by ELISA (borderline 45.92; reactive 91.83). SARS-CoV-2 RT-PCR confirmed patients are shown in red. Horizontal bars depict the median. IgG: Immunoglobulin G, NP: nucleocapsid protein, RBD: receptor binding domain, OD: optical density, S/CO: signal-to-cut-off ratio, IU: international unit. < 0.0001 ****, 0.0001 to 0.001 ***, 0.001 to 0.01 **, 0.01 to 0.05 *, ≥ 0.05, not significant, ns.

Sera were not reactive for anti-SARS-CoV-2-NIgG except for four cases of PCR-confirmed SARS-CoV-2 infection (**Figure 1D**, indicated in red) and one patient of the unvaccinated control group. This patient did not show antibodies against the spike antigens and did not test positive in IGRA. This could be explained by a test artifact or by partial immune response after infection. SARS-CoV-2 specific T-cells, were present three weeks after the second vaccination in 21/31 patients (67.74%; 95%CI: 48.53-82.68) (**Figure 1E**). IGRA correlated positively and significantly (p<0.001) with anti-SARS-CoV-2-S1 IgG (**eFigure 3 in Supplements**), confirming a specific response of both B and T cells to Tozinameran.

Antibody levels of vaccinated patients on dialysis ∼3-4 weeks after the second dose were significantly (p<0.01) lower than in the vaccinated control cohort sampled four weeks after the second dose for all four test systems (**Figure 1B, C, eFigure 4, eFigure 5 in Supplements**). In line with this, percentage of patients with antibody detection above the threshold was higher for the vaccinated control cohort than for patients on dialysis. For fS and RBD antigens these rates differ significantly, with 77.14% (95%CI: 59.44-88.95) and 71.43% (95%CI: 53.48-84.76) for patients on dialysis versus 95.45% (95%CI: 83.29-99.21) and 93.18% (95%CI: 80.29-98.22) for controls, respectively (both p<0.05). For the S1 antigen and the sNT, differences between cohorts were similar, although not statistically significant with 77.14% (95%CI: 59.44-88.95) and 77.78% (95%CI: 60.42-89.28) seropositivity for patients on dialysis versus 90.91% (95%CI: 77.42-97.05) and 87.80% (95%CI: 72.99-95.42) for controls, respectively (p=0.09 for S1 antigen and p=0.39 for sNT).

T-cell response assessed by IGRA turned out positive at significantly lower ratios with 67.74% (95%CI: 48.53-82.68) for patients on dialysis compared to 93.33% (95%CI: 76.49-98.84) of the controls (p=0.03). Corresponding interferon gamma levels in the dialysis cohort compared to the vaccinated control cohort was also significantly lower (p<0.05, **Figure 1E**). Serologic non-responders showed negative results for SARS-CoV-2 specific T-cell responses.

## Discussion

We demonstrate a high percentage of SARS-CoV-2 specific seroreactivity after mRNA vaccination with Tozinameran in patients on dialysis. The observed response was significantly lower compared to control patients of similar age not on dialysis as well as otherwise reported patients older than 75 years of age [3]. Ultimately, around 10% remained with no detectable response to vaccination in the SARS-CoV-2-S1 IgG and IgA ELISA.

In line with our study on COVID-19 vaccine, impaired vaccine immunogenicity in patients on chronic dialysis has been reported for those with viruses such as influenza and hepatitis B [7 8]. The decreased number of B and T cells in patients with kidney failure undergoing chronic dialysis might contribute to the differences between patients with KF and without dialysis [9-11]. Moreover, impaired clearance of uremic toxins, systemic inflammation, and malnutrition might contribute to lower immunogenicity (reviewed in [12]). These factors need to be addressed in further studies. However, since patients with KF were excluded from previous COVID-19 vaccine trials, our data present an important report of immunogenicity of COVID-19 vaccines in patients on dialysis.

A recent study from Israel, where 1.2M people have been vaccinated, demonstrated that infection risks vary with patient-specific characteristics [13]. These included age and comorbidities, e.g. immunosuppression and type 2 diabetes [13], as risk for a lower immune response. Consistent with that, less robust immune responses were observed in solid organ transplant recipients after one dose of mRNA vaccines [14]. Of note, in our dialysis cohort, three patients developed COVID-19 after first vaccination, of whom one died of COVID-19, underlining the need for ongoing non-pharmaceutical intervention after vaccinations, especially in these patients

Limitations of our studies include the small cohort size and the use of only one mRNA vaccine. Therefore, further risk factors contributing to a negative immune response remain to be defined. Prospective studies with other vaccines against COVID-19, e.g., viral vector–based vaccines, will help to elucidate the efficacy of different vaccines in these patients. Moreover, antibody titer persistence in patients on dialysis might differ from otherwise healthy persons, which should be addressed in longitudinal observations.

## Conclusion

Although our data contrast the robust response rate to Tozinameran in the elderly (≥75 years) [3], the overall high percentage of a response three weeks after the second dose, should encourage rapid vaccination of this vulnerable group. Patients without detectable immune response might need attention and potentially a booster vaccination. Our data demonstrate the urgent need for more detailed studies of the immune response to SARS-CoV-2 vaccines in populations with underlying clinical conditions.

## Data Availability

All data are given within the manuscript.

## Article Information

## Additional Contributions

We are grateful to the patients who participated in this trial.

## Author Affiliations

Department of Nephrology and Intensive Care, Charité-Universitätsmedizin Berlin, corporate member of Freie Universität Berlin and Humboldt-Universität zu Berlin, Berlin, Germany (Schrezenmeier, Weber, Stefanski, Budde, Halleck, Choi). Institute of Virology, Charité-Universitätsmedizin Berlin, Humboldt-Universität zu Berlin, Berlin Institute of Health, and German Centre for Infection Research (DZIF), Partner Site Charité, Berlin, Germany. (Bergfeld, Schwarz, Corman). Department of Infectious Diseases and Respiratory Medicine, Charité-Universitätsmedizin Berlin, corporate member of Freie Universität Berlin and Humboldt-Universität zu Berlin, Berlin, Germany (Hillus, Tober-Lau, Sander, Kurth). Nierenzentrum Koethen, Koethen, Germany (Lippert). Hausarztpraxis am Agaplesion Bethanien Sophienhaus, Berlin, Germany (Landgraf). Institute of Laboratory Medicine, Clinical Chemistry and Pathobiochemistry, Charité – Universita□tsmedizin Berlin, corporate member of Freie Universita□t Berlin and Humboldt-Universita□t zu Berlin, and Labor Berlin – Charité Vivantes GmbH, Berlin, Germany (Kappert). Department for General, Visceral and Vascular Surgery, Charité-Universitätsmedizin Berlin, corporate member of Freie Universität Berlin and Humboldt-Universität zu Berlin, Berlin, Germany (Sattler, Kotsch). Department of Rheumatology and Clinical Immunology, Charité-Universitätsmedizin Berlin, corporate member of Freie Universität Berlin and Humboldt-Universität zu Berlin, Berlin, Germany (Doerner). Department of Tropical Medicine, Bernhard-Nocht Institute for Tropical Medicine, Hamburg, Germany (Kurth)

