## Supplements for "Immunogenicity of COVID-19 Tozinameran Vaccination in Patients on Chronic Dialysis"

| **Table of Contents** |  |  |
| --- | --- | --- |
| Patient and clinical characteristics |  | Page 2 |
| Additional eFigure 1. Study Flow Diagram |  | Page 3 |
| Ethical Statement |  | Page 3 |
| Detailed Methods |  | Page 4 |
| Additional eTable 1. Positive Outcome in the different test systems |  | Page 7 |
| Additional eFigure 2. SARS-CoV-2 IgA responses in vaccinated and unvaccinated dialysis-patients. |  | Page 9 |
| Additional eFigure 3: Correlation of anti-SARS-CoV-2 spike S1 IgG and IGRA in vaccinated dialysis patients |  | Page 9 |
| Additional eFigure 4: Detection of anti-SARS-CoV-2 S1 IgG in vaccinated dialysis patients and non-dialysis patients by SeraSpot anti-SARS-CoV-2 IgG Assay. |  | Page 10 |
| Additional eFigure 5: Detection of anti-SARS-CoV-2 spike full IgG in vaccinated dialysis patients and non-dialysis patients by SeraSpot anti-SARS-CoV-2 IgG Assay. |  | Page 10 |
| Additional eFigure 6: IFN-γ concentration after mitogen stimulation. |  | Page 11 |

**Patient and clinical characteristics**

43 patients with chronic kidney disease (CKD) stage 5 were initially vaccinated with Tozinameran (BNT162b2 BioNTech/Pfizer). Three out of 43 patients developed SARS-CoV-2 infection at 11, 14 and 16 days after the first vaccination, of whom one patient died from COVID-19 infection 19 days after the first vaccination. One patient died from sepsis 37 days after first vaccination (unrelated to vaccination and COVID-19) and three patients missed at least one follow-up measurement, thus the remaining 36 patients were included in the study. Of these 34 patients were on hemodialysis and 2 on peritoneal dialysis. We analyzed samples from two time points: 1^st^ samples were taken on average 7.8 days (range 7-13) after the second dose and the 2^nd^ samples were taken on average 21.2 days (range 20-26) after second dose, which was given 21 days apart. In addition to serum samples, full blood (Li-Heparin) for T-cell assays was taken at the 2^nd^ time point. The study flow diagram is shown in Figure S1.

For controls, we recruited 18 patients with CKD stage 5 on hemodialysis, 2 of them were reported to have RT-PCR confirmed COVID-19 in the past (2 month and 3 months before sampling). Median age was 63 years (IQR:45.25, 74.5) with a female to male ratio of 6:12.

We further investigated subjects from two ongoing observational cohort studies on immunogenicity of SARS-CoV-2 vaccines. 44 subjects from these studies were selected as controls for the dialysis patients by univariate matching for age. Median age was 80 years (IQR: 75.8, 82.2) and 30 subjects (68%) were female. None of these subjects were on hemodialysis. All subjects received two doses of the Tozinameran 21 days apart and were sampled on average 26.6 days (range 25-34) after the 2^nd^ vaccination.

**eFigure 1. Inclusion Diagram dialysis patients**

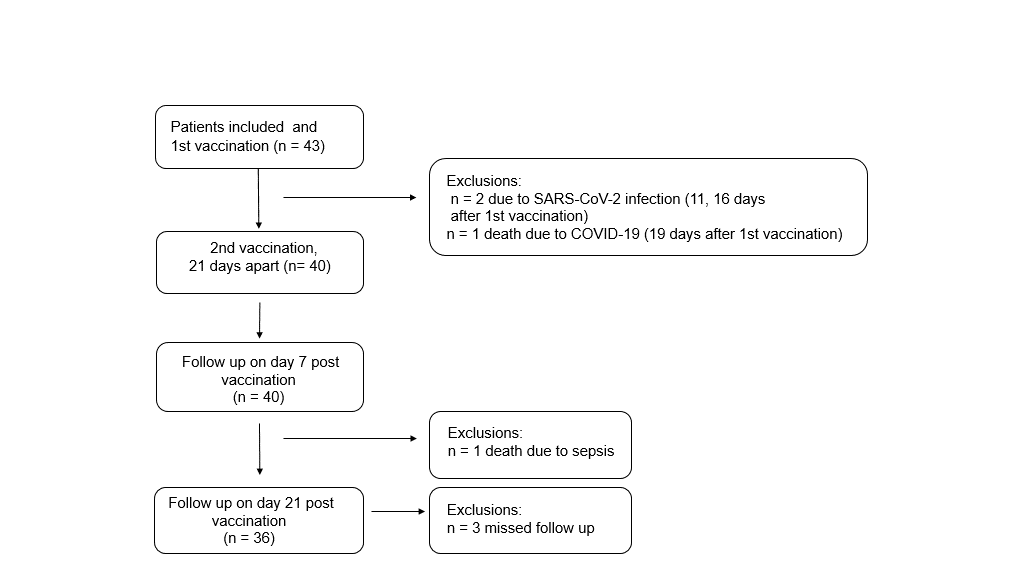

**Ethical statement**

This study was approved by the local institutional review board of the ethics committee of Charité-Universitätsmedizin Berlin, Germany (approval number EA4/188/20) and by the local ethics committee of Sachsen-Anhalt (EA7/21). The testing of non-dialysis control subjects is part of ongoing studies on SARS-CoV-2 infection and immunogenicity of COVID-19 vaccines in elderly people and healthcare workers under Charité ethical review board file numbers EA1/068/20, EA4/244/20, and EA4/245/20. Written informed consent was obtained from all patients and healthy volunteers before inclusion in the studies.

**Detailed methods**

**Serological testing for SARS-CoV-2 antibodies**

To test for SARS-CoV-2 specific antibodies, we performed anti-SARS-CoV-2 S1 IgG and IgA ELISAs according to the manufacturer’s instructions (Euroimmun Medizinische Labordiagnostika AG, Lübeck, Germany). Serum samples were analyzed at a 1:101 dilution using the fully automated Euroimmun Analyzer I (Euroimmun Medizinische Labordiagnostika AG). Optical density (OD) ratios above 1.1 were considered reactive for IgG and IgA.

To discriminate from previous SARS-CoV-2 infection and to confirm reactivity in the Euroimmun S1-ELISA, a solid phase immunoassay (SeraSpot®Anti-SARS-CoV-2 IgG, Seramun Diagnostica GmbH, Heidesee, Germany) for the detection of antibodies present in isolated samples was applied. This assay is based on the use of four recombinant SARS-CoV-2 proteins (complete spike, S1 domain, RBD and nucleocapsid protein) as capture antigens. These and test specific controls are printed in an array arrangement on the bottom of each of the well. Bound antibodies from the patient sera are detected by use of horseradish peroxidase-(HRP)-labeled antibodies against human IgG. Color intensity at the site of formed immune complexes (pale to dark blue) correlates to the antibody concentration. The SpotSight®plate scanner was used for measurements. Results were calculated and as normalized signal-to-cutoff (S/CO) ratios by dividing the observed signal strength of a specific spot by that of an internal cutoff control. Samples with an S/CO ratio of ≥ 1.0 are defined by the manufacturer as positive.

**Interferon-gamma release assay (IGRA)**

We applied a commercially available interferon-gamma release assay (IGRA) for assessment of IFN-γ release of SARS-CoV-2-specific T-cells. (Euroimmun Medizinische Labordiagnostika AG, Lübeck, Germany). Here, in parallel 0.5 mL freshly collected lithium-heparin blood (~ 4 hours after blood sampling) was stimulated with a SARS-CoV-2 peptide pool from the spike domain, 0.5 mL of blood was stimulated with mitogen as a positive control (eFigure S6), and 0.5 mL of blood in a blank as a negative control. After 24 hours of incubation (at 37°C) IFN-γ concentration in the plasma fraction of all three stimulation tubes was measured by ELISA. IFN-γ response in the blank served as a measure of patient-individual background IFN-γ activity and was subtracted from IFN-γ response in the stimulation tubes. For analyses of IGRA outcome, we defined an arbitrary cutoff by using average IFN-γ activity (9.8 mIU/mL) determined in the 16 SARS-CoV-2 IgG antibody negative none-vaccinated dialysis control multiplied with 5 as the threshold (45.92 mIU/mL) for borderline IGRA reactive and multiplied by 10 for the threshold (91.83 mIU/mL) for IGRA positive. Using this cutoff, we found 2 out of 2 infected but unvaccinated dialysis subjects to be classified positive in the IGRA, but none of the unvaccinated dialysis controls. Only samples showing an IFN-γ activity of at least 100 mIU/mL in the mitogen stimulated tube were further analyzed.

**Surrogate SARS-CoV-2 neutralization test (cPass)**

To detected neutralizing activity in serum samples, an ELISA-based SARS-CoV-2 surrogate virus neutralization assay (sVNT) was used according to the manufacturer’s instructions (cPass Assay, Medac, Wedel, Germany). Briefly, serum samples, positive and negative controls were diluted 1:10 with sample dilution buffer, mixed 1:1 with HRP-RBD solution and incubated at 37°C for 30 minutes. 100 µl of each sample was added to the hACE2 coated plate and incubation at 37°C for 15 minutes. After a washing step, TMB solution was added, and the plate was incubated in the dark at room temperature for 15 minutes. Then, 50 µl stop solution was added per well and the optical density at 450 nm was measured. For calculation of the inhibition in % following formulation was used: Inhibition score (%) = (1 - (OD value_sample_/OD value_negative control_) x 100%). The samples were considered negative at an inhibition of <30 % and positive at an inhibition of ≥ 30 %.

**Statistics**

Statistical analyses were performed using GraphPad Prism Version 9. Differences between groups were assessed by Mann‐Whitney U test (two groups) or Kruskal Wallis test (more than two groups). Correlations were calculated using Spearman's rank coefficient. P values less than 0.05 were considered significant. Qualitative outcomes of different cohorts of vaccinees were assessed using Chi square test with Yates' correction. All 95% confidence intervals for proportions were calculated using the Wilson procedure with a correction for continuity.

**eTable 1.** Positive outcome in the different test systems. Percentage with positive result, (95% CI), [no. pos/ no. tested] per cohort*.

| **Cohort** | **Vaccinated**  **dialysis patients** | **Control**  **dialysis patients** | **Control non-dialysis patients** | **Comparison of qualitative outcomesbetween vaccinated groups** |
| --- | --- | --- | --- | --- |
| **IgG ELISA**  **(1^st^ sampling)** | 55.56 (38.29-71.67)  [20/36] | 0.00 (0.00-24.07)  [0/16] | Not tested | Not tested |
| **IgG ELISA**  **(2^nd^ sampling)** | 88.89 (73.00-96.38)  [32/36] | Not tested | Not tested | Not tested |
| **IgA ELISA S1**  **(1^st^ sampling)** | 61.11 (43.53-76.37)  [22/36] | 12.25 (2.20-39.59)  [2/16] | Not tested | Not tested |
| **IgA ELISA S1**  **(2^nd^ sampling)** | 91.67 (76.41-97.82)  [33/36] | Not tested | Not tested | Not tested |
| **NP spot array** | 2.86 (0.15-16.62)  [1/35] | 6.25 (0.33-32.29)  [1/16] | 0.00 (0.00-10.00)  [0/44] | Not tested |
| **RBD spot array** | 77.14 (59.44-88.95)  [27/35] | 0.00 (0.00-24.07)  [0/16] | 95.45 (83.29-99.21)  [42/44] | Yates' chi-square = 4.37  Yates' p-value = 0.04 |
| **S1 spot array** | 74.29 (56.43-86.90)  [26/35] | 0.00 (0.00-24.07)  [0/16] | 90.91 (77.42-97.05)  [40/44] | Yates' chi-square = 2.80  Yates' p-value = 0.09 |
| **Full Spike spot array** | 71.43 (53.48-84.76)  [25/35] | 0.00 (0.00-24.07)  [0/16] | 93.18 (80.29-98.22)  [41/44] | Yates' chi-square = 5.22  Yates' p-value = 0.02 |
| **RBD/ACE2 Inhibition** | 77.78 (60.42-89.28)  [28/36] | 0.00 (0.00-43.91)  [0/7] | 87.80 (72.99-95.42)  [36/41] | Yates' chi-square = 0.75  Yates' p-value = 0.39 |
| **IGRA**  **T-cell reactive** | 67.74 (48.53-82.68)  [21/31] | 0.00 (0.00-24.07)  [0/16] | 93.33 (76.49-98.84)  [28/30] | Yates' chi-square = 4.80  Yates' p-value = 0.03 |

*** Excluding the SARS-CoV-2 RT-PCR- positive patients.

CI: Confidence Interval

RBD: Receptor-Binding Domain

ACE2: Angiotensin-Converting Enzyme 2

IGRA: Interferon Gamma Release Assay

**eFigure 2**

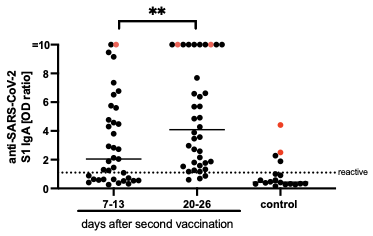

**eFigure 2: SARS-CoV-2 IgA responses in vaccinated and non-vaccinated dialysis-patients.** Anti-SARS-CoV-2 S1 IgA were measured in serum of dialysis patients on day 7-13 (n=37) and day 20-26 (n=38) after a second vaccination with BNT162b2 and non-vaccinated dialysis patients (n=18) utilizing a EUROIMMUN ELISA. SARS-CoV-2 RT-PCR-confirmed patients are shown in red. Horizontal bars depicted the median. < 0.0001 ****, 0.0001 to 0.001 ***, 0.001 to 0.01 **, 0.01 to 0.05 *, ≥ 0.05, Not significant, ns.

**eFigure 3:**

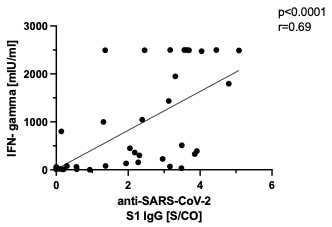

**eFigure 3: Correlation of anti-SARS-CoV-2 S1 IgG with IFN-γ concentration after S1 stimulation.** Anti-SARS-CoV-2 S1 IgG were detected in serum of dialysis patients and controls 20-26 days after the second vaccination with Tozinameran by using the SeraSpot anti-SARS-CoV-2 assay and correlated with interferon-γ release after S1 stimulation.

**eFigure 4**

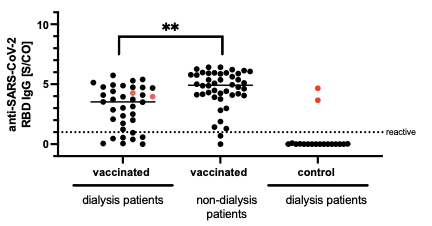

**eFigure 4: Detection of anti-SARS-CoV-2 RBD IgG in vaccinated dialysis patients and non-dialysis patients by SeraSpot anti-SARS-CoV-2 IgG Assay.** Anti-SARS-CoV-2 S1 IgG were detected in serum of dialysis patients 20-26 days after the second vaccination with Tozinameran (n=36) and unvaccinated dialysis patients (n=18) which were compared with vaccinated non-dialysis patients (n=44) by using the SeraSpot anti-SARS-CoV-2 assay. One vaccinated dialysis patient was excluded as the internal assay control failed. SARS-CoV-2 RT-PCR-confirmed patients are shown in red. Horizontal bars depicted the median. S/CO: signal-to-cutoff ratio. < 0.0001 ****, 0.0001 to 0.001 ***, 0.001 to 0.01 **, 0.01 to 0.05 *, ≥ 0.05, not significant, ns.

**eFigure 5**

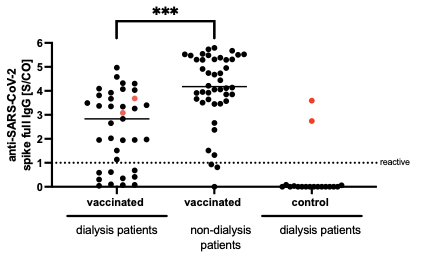

**eFigure 5: Detection of anti-SARS-CoV-2 full spike IgG in vaccinated dialysis patients and non-dialysis patients by SeraSpot anti-SARS-CoV-2 IgG Assay.** Anti-SARS-CoV-2 spike full IgG were detected in serum of dialysis patients 20-26 days after the second vaccination with Tozinameran (n=36) and unvaccinated dialysis patients (n=18) which were compared with vaccinated non-dialysis patients (n=44) by using the SeraSpot anti-SARS-CoV-2 assay. One vaccinated dialysis patient was excluded as the internal assay control failed. SARS-CoV-2 RT-PCR-confirmed patients are shown in red. Horizontal bars depicted medians. S/CO: signal-to-cutoff ratio. < 0.0001 ****, 0.0001 to 0.001 ***, 0.001 to 0.01 **, 0.01 to 0.05 *, ≥ 0.05, not significant, ns.

**eFigure 6**

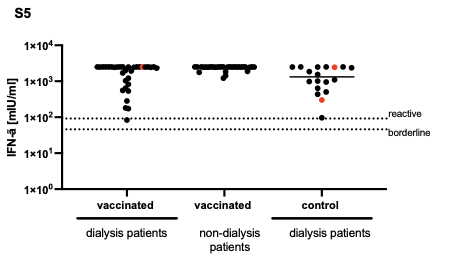

**eFigure 6: IFN-γ concentration after mitogen stimulation.** Whole blood from vaccinated dialysis patients (n=33), unvaccinated dialysis patients (n=18) and non-dialysis patients (n=30) was stimulated ex vivo for 24 h with mitogen and IFN-γ concentration in the supernatant was determined by an interferon-γ release assay (IGRA). Horizontal bars depict the median.
